# Safety and efficacy of three-cavity clearance in the management of cryptoglandular perianal abscess: A multicentre randomized controlled study

**DOI:** 10.1101/2020.04.10.20061309

**Authors:** Xin-Yi Zhang, Cheng Chen, Ping Wei, Yu-Yao Wang, Li-Jiang Ji, Qiong Zhang, Chun-Xia Zhang, Hei-Ying Jin

**Affiliations:** Department of Colorectal Surgery, Second Affiliated Hospital of Nanjing University of Chinese Medicine, Nanjing 210017, Jiangsu Province, China; Rugao Hospital of Traditional Chinese Medicine, Rugao 226500, Jiangsu Province, China; Lianyungang Hospital of Traditional Chinese Medicine, Lianyungang 222001, Jiangsu Province, China; Changshu Hospital of Traditional Chinese Medicine, Changshu 215500, Jiangsu Province, China; Shuyang Hospital of Traditional Chinese Medicine, Shuyang 223699, Jiangsu Province, China

**Author notes:** These authors equally contributed to the work (Xin-Yi Zhang, Cheng Chen, Ping Wei). **Corresponding author:** Hei-Ying Jin,M.D., Department of colorectal surgery, The Second Affiliated Hospital of Nanjing University of Chinese Medicine, 23 Nanhu Road, Jianye District, Nanjing 210017, Jiangsu Province, China. **Financial Disclosure:** None reported. **Funding/Support:** None received. Quick shot of distinction presentation at the meeting of The American Society of Colon and Rectal Surgeons, Boston, MA, June 6-10, 2020. IF the manuscript is provisionally accepted, a VIDEO ABSTRACT will be required prior to final acceptance.

**Keywords:** perianal abscess, three-cavity clearance, simple incision and drainage, multicentre randomized controlled study

## Abstract

**OBJECTIVE:** To evaluate the safety and efficacy of three-cavity clearance in the management of cryptoglandular perianal abscess.

**METHOD:** This was a multicentre randomized controlled study. The study was designed and approved by the ethics committee of the Second Affiliated Hospital of Nanjing University of Traditional Chinese Medicine. The study was registered in the Chinese Clinical Trial Register centre (ChiCTR1800016958).Patients with cryptoglandular perianal abscess in 5 Hospitals from Sept.2018 to Sept. 2019 were included.The anal fistula rate, anal incontinence, abscess recurrence, success rate, postoperative pain, wound healing time, and hospitalization duration were compared.

**RESULTS:** Total 334 patients were enrolled in the study, who were 162 in the three-cavity clearance group and 172 in the control group. The anal fistula rate and abscess recurrence rate were 6.2% and 1.9% in the three-cavity clearance group (P=0.001) and 18.0% and 8.1% in the control group (P=0.009). No patients experienced fecal incontinence. The success rate in the three-cavity clearance group was 92.0% and that in the control group was 73.8% (P=0.00001). The postoperative pain on day 3 was lower in the three-cavity clearance group than that in the control group (P=0.002). The hospitalization duration was 9.0±5.4 days in the three-cavity clearance group and 10.4±6.1days in the control group (P=0.049). The wound healing time was 27.1±16.4 days in the three-cavity clearance group and 28.2±14.1 days in the control group (P=0.764).

**CONCLUSIONS:** This randomized controlled study showed that three-cavity clearance is a safe and effective management of cryptoglandular perianal abscess.

## Introduction

Perianal abscess is a common perianal infectious disease, with an incidence of as high as 2-10 cases per 10000 inhabitants per year according to a German guideline, and is more common in young men aged 20–40 years [1–3]. It is believed that anal gland infection are thought to be responsible for 90% of cases, and timely operation is required for treatment [4, 5]. The most common surgical procedure is simple incision and drainage; however, it is associated with high rates of postoperative abscess recurrence or anal fistula formation [6]. To reduce the postoperative anal fistula rate, some surgeons directly open the suspected fistula or use the cutting seton procedure at the same time as abscess incision and drainage, which is called ‘radical abscess incision’. However, there exists a high risk of postoperative fecal incontinence caused by injury to the anal sphincter [7, 8]. Further, most surgeons believe that inaccurate detection of the fistula and internal opening during acute infection results in an increased risk of postoperative false pathway and fecal incontinence [9–11].

Thereby, we designed a new procedure based on the physiological and pathological mechanisms of perianal abscess, which is called ‘three-cavity clearance (TCC)’. According to the route of perianal abscess spread, the anorectal space was divided into three cavities: the submucosal cavity between the mucosa and the internal sphincter, the intersphincteric cavity between the internal and external sphincter, and the cavity outside the external sphincter (including the ischiorectal, presacral, and pelvirectal cavities) (Figure 1). We hypothesized that opening all potentially infected cavities and performing full drainage might reduce the postoperative anal fistula rate and cure the disease without damaging the anal function [12]. In a previous prospective cohort study [13] and a retrospective case-control study [14], the fistula formation rates in patients who underwent TCC were 6% and 13%, respectively, which were significantly lower than the rates in patients who underwent simple incision and drainage (34% and 39.1%, respectively). In both studies, no anal incontinence occurred in the two groups. In addition to the data collection and analysis of the clinical case study, we also used three-dimensional endorectal ultrasonography for preoperative and postoperative evaluations, and there was no residual pus cavity postoperatively (Figure 2). To further confirm whether TCC is a safe and effective method for the treatment of cryptoglandular perianal abscess, we performed the present multicentre, randomized, controlled study.

**FIGURE 1.**
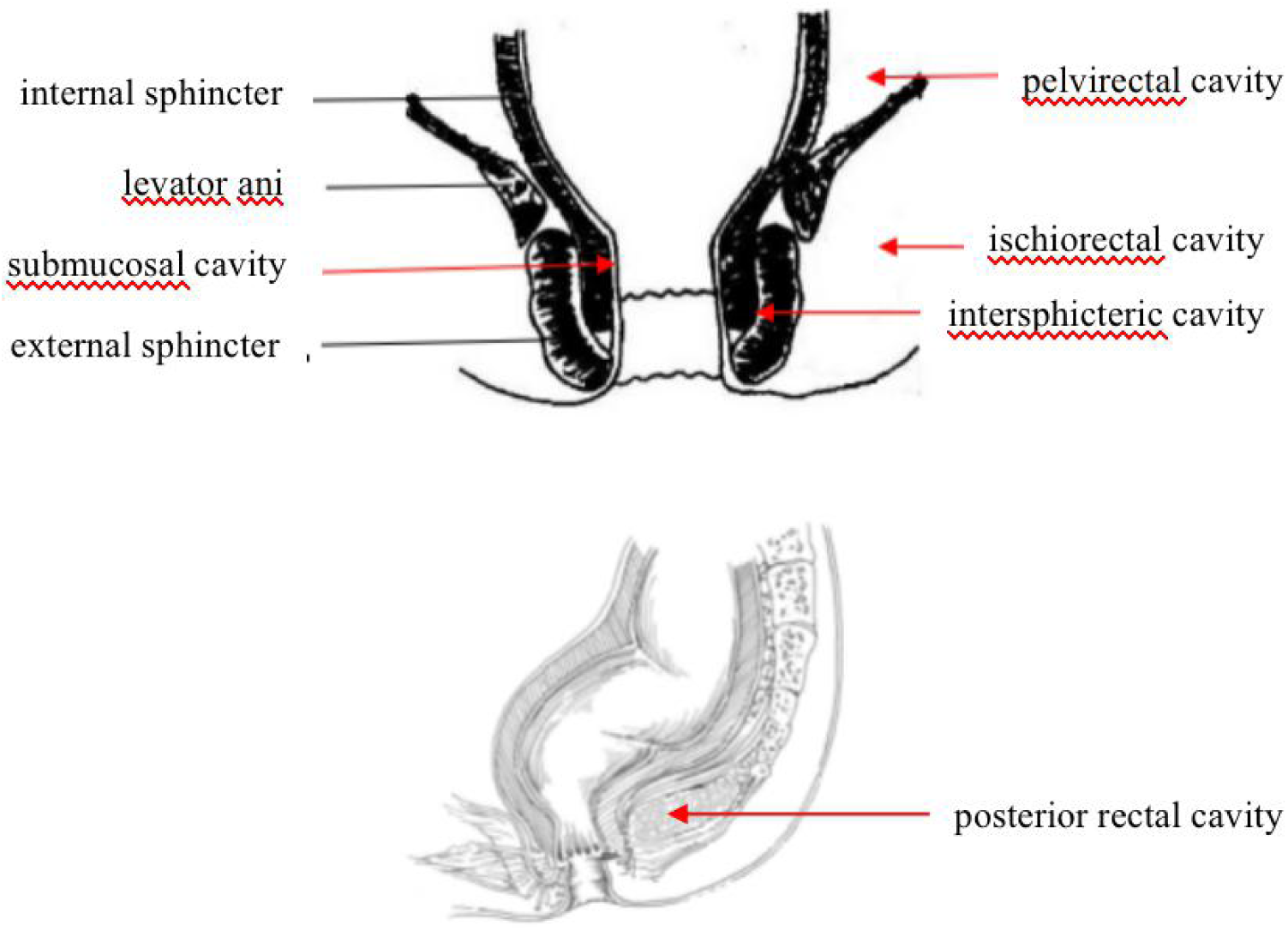
Diagram of the three cavities

**FIGURE 2.**
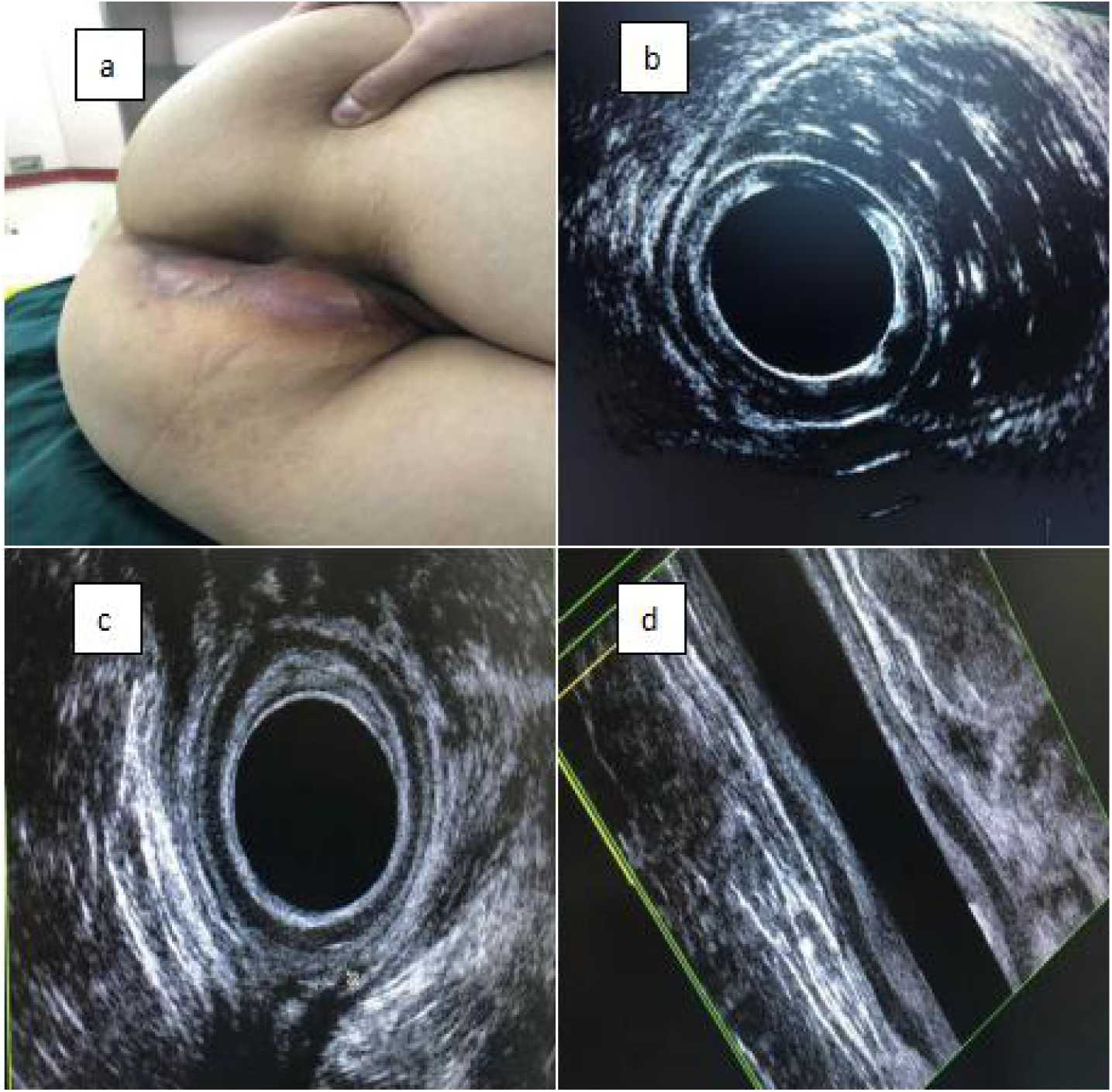
Three-dimensional endorectal ultrasonography(3D-EAUS). a, b refers to the appearance of ischiorectal abscess under preoperative 3D-EAUS; c and d were seen under postoperative 3D-EAUS, the internal and external sphincter remained intact, and there was no residual pus cavity.

### Patients and methods

#### 1. General information

Patients with perianal abscess who were treated in the Department of Colorectal Surgery of the Second Affiliated Hospital of Nanjing University of Traditional Chinese Medicine (TCM), Rugao Hospital of TCM, Lianyungang Hospital of TCM, Changshu Hospital of TCM, and Shuyang Hospital of TCM from September 2018 to September 2019 were included in this study. A multicentre, randomized, parallel, and controlled method was adopted. A total of 334 patients were enrolled in the study, who were divided into the TCC group (n=162) and the simple incision and drainage group (control group; n=172) through the random envelope method. The inclusion criteria were as follows: presence of perianal abscess on the basis of the diagnostic criteria in the Practice Parameters for the Management of Perianal Abscess and Fistula-in-Ano (2011, USA) [15], and a diagnosis of perianal abscess based on endorectal ultrasonography (EUS) or magnetic resonance imaging (MRI) findings. The exclusion criteria were as follows: (1) complications such as tuberculosis, Crohn’s disease, colorectal tumour, and other specific perianal abscess; (2) immunodeficiency diseases; (3) pregnancy and lactation in female patients; and (4) history of previous abscess surgery with the occurrence of anal dysfunction and poor compliance. The study was approved by the ethics committee of the Second Affiliated Hospital of Nanjing University of TCM and registered in the Chinese Clinical Trial Register centre (ChiCTR1800016958). All patients provided informed consent.

#### 2. Operative procedures

TCC was performed according to the method reported in our previous study [12–14]. The method of simple incision and drainage was as previously described [15].

Preoperative preparation and postoperative management were performed routinely.

#### 3. Observation indices

Through clinical observations, outpatient observations, and telephone follow-up, the postoperative anal fistula rate, anal incontinence (Wexner score), abscess recurrence, success rate, postoperative pain visual analogue scale (VAS) score[16], wound healing time, and hospitalization duration were compared between the two groups. The diagnosis of postoperative anal fistula was based on the Practice Parameters for the Management of Perianal Abscess and Fistula-in-Ano (2011, USA) [15]. Anal incontinence was assessed using the Wexner score [17], with 0 indicating normal and 20 indicating complete incontinence. The repeated formation of abscess at the same site or failure to cure the same abscess for >2 months was defined as abscess recurrence[28]. Success was defined as the absence of anal fistula, abscess recurrence, and anal dysfunction after the operation. The VAS score was used to evaluate postoperative pain in all patients on the day of the operation and on days 1, 2, 3, and 7 after the operation, with a score of 0 indicating no pain and 10 indicating extreme pain. The hospitalization duration was defined as the time from the first day of admission to the day of discharge. The wound healing time was calculated from the first day after surgery to the day when the wound was completely epithelialized.

#### 4. Statistical methods

SPSS23.0 software (SPSS Inc., Chicago, IL, USA) was used for the analysis. Measurement data are presented as means with standard deviation, and one-way analysis of variance was used for comparison among groups. The least significant difference method was used for pairwise comparison. Enumeration data are expressed as number of cases or percentage, and the chi-square test was used for comparison among groups. P<0.05 indicated that the difference was statistically significant.

## Results

A total of 334 patients were included in this study, comprising 270 men and 64 women. Statistical analysis showed no significant difference in general information including age, gender, and abscess site between the two groups (P>0.05, Table 1). All patients were followed up with EUS or MRI examination in the outpatient department, with an average follow-up period of 8.5±3.2 months.

**TABLE 1.** General information of patients

|  | TCC group<br>(n=162) | Control group<br>(n=172) | TCC group vs. control<br>group |
| --- | --- | --- | --- |
|  |  |  | P-value |
| Age, years | 41.9±13.3 | 39.1±13.1 | 0.082 |
| Sex |  |  |  |
| Male | 130 | 140 | 0.789 |
| Female | 32 | 32 |  |
| Abscess site |  |  |  |
| Perianal subcutaneous<br>abscess | 52 | 78 | 0.067 |
| Intersphincter abscess | 58 | 46 |  |
| Ischiorectal abscess | 21 | 20 |  |
| Pelvirectal abscess | 11 | 5 |  |
| Retrorectal abscess | 20 | 23 |  |

### 1. Hospitalization duration

The hospitalization duration in the control group ranged from 3 to 29 days (average, 10.4±6.1 days). The hospitalization duration in the TCC group ranged from 5 to 28 days (average, 9.0±5.4 days). The difference in hospitalization duration between the two groups was statistically significant (P=0.049, Table 2).

**TABLE 2.** Outcomes of the two groups after surgery

|  | TCC group<br>(n=162) | Control group<br>(n=172) | P-value |
| --- | --- | --- | --- |
|  |  |  | TCC group vs.<br>control group |
| Wound-healing time, days | 27.1±16.4 | 28.2±14.1 | 0.764 |
| Hospitalization duration,<br>days | 9.0±5.4 | 10.4±6.1 | 0.049 |
| Fistula rate, n (%) | 10 (6.2) | 31 (18.0) | 0.001 |
| Recurrence, n (%) | 3 (1.9) | 14 (8.1) | 0.009 |
| Success rate, n (%) | 149 (92.0) | 127 (73.8) | 0.00001 |
| Anal incontinence, n (%) | 0 (0.0) | 0 (0.0) |  |

### 2. Wound healing time

The healing time in the control group ranged from 7 to 113 days (average, 28.2±14.1 days). The healing time in the TCC group ranged from 16 to 123 days (average, 27.1±16.4 days). The difference in wound healing time between the two groups was no statistically significant (P=0.764, Table 2).

### 3. Postoperative pain VAS score

The postoperative VAS score in the TCC group on day 3 was 2.4±1.4 points, which was lower than that in the control group (2.9±1.5 points), and the difference was statistically significant (P=0.002, Table 3). No statistically significant difference was found in the postoperative VAS scores of the two groups on days 0 (operative day), 1, 2, and 7 (P=0.949, P=0.114, P=0.178, P=0.205; Table 3).

**TABLE 3.** Postoperative visual analogue scale score

|  | TCC group<br>(n=162) | Control group<br>(n=172) | P-value |
| --- | --- | --- | --- |
|  |  |  | TCC group vs.<br>control group |
| Day 0 (operative day) | 2.4±1.6 | 2.3±1.2 | 0.949 |
| Day 1 | 3.1±1.5 | 3.4±1.5 | 0.114 |
| Day 2 | 2.9±1.6 | 3.3±1.6 | 0.178 |
| Day 3 | 2.4±1.4 | 2.9±1.5 | 0.002 |
| Day 7 | 1.9±1.2 | 2.2±1.1 | 0.205 |

### 4. Fistula formation

Anal fistula formed in 31 patients in the control group (23 men and 8 women), including 2 patients with perianal subcutaneous abscess, 7 patients with intersphincter abscess, 10 patients with ischiorectal abscess, 9 patients with presacral abscess, and 3 patients with pelvirectal abscess. There were 10 patients with anal fistula in the TCC group (8 men and 2 women), including 1 patient with perianal subcutaneous abscess, 1 patient with intersphincter abscess, 3 patients with ischiorectal abscess, 4 patients with presacral abscess, and 1 patient with pelvirectal abscess (Table 4). The anal fistula formation rate (18.0%) in the control group was significantly higher than that in the TCC group (6.2%) (P=0.001, Table 2).

**TABLE 4.** Anal fistula formation

|  |  | TCC group | Control group |
| --- | --- | --- | --- |
| n |  | 10 | 31 |
| Sex | Male | 8 | 23 |
|  | Female | 2 | 8 |
| Abscess site | Perianal subcutaneous abscess | 1 | 2 |
|  | Intersphincter abscess | 1 | 7 |
|  | Ischiorectal abscess | 3 | 10 |
|  | Retrorectal abscess | 4 | 9 |
|  | Pelvirectal abscess | 1 | 3 |

### 5. Recurrence

There were 14 patients with abscess recurrence in control group (10 men and 4 women), including 2 patients with perianal subcutaneous abscess, 7 patients with intersphincter abscess, 3 patients with ischiorectal abscess, 1 patient with presacral abscess, and 1 patient with pelvirectal abscess. Three patients in the TCC group had recurrent abscess after surgery (2 men and 1 woman), including 1 patient with perianal subcutaneous abscess, 1 patient with intersphincter abscess, and 1 patient with presacral abscess (Table 5). The abscess recurrence rate in the TCC group was 1.9%, which was lower than that in the control group (8.1%), and the difference was statistically significant (P=0.009, Table 2).

**TABLE 5.** Abscess recurrence

|  |  | TCC group | Control group |
| --- | --- | --- | --- |
| n |  | 3 | 14 |
| Sex | Male | 2 | 10 |
|  | Female | 1 | 4 |
| Abscess site | Perianal subcutaneous abscess | 1 | 2 |
|  | Intersphincter abscess | 1 | 7 |
|  | Ischiorectal abscess | 0 | 3 |
|  | Retrorectal abscess | 1 | 1 |
|  | Pelvirectal abscess | 0 | 1 |

### 6. Anal function

During the follow-up, there were no cases of fecal incontinence in the two groups.

### 7. Success rate

The success rate of TCC group was 92.0% and that of control group was 73.8%, with a statistically significant difference (P=0.00001, Table 2).

## Discussion

After conventional simple incision and drainage, the anal fistula rate is about 7–66% and the recurrence rate is 4–31% [18], which may be related to poor incision and drainage, improper treatment of the internal opening, and complicated infection between sphincters [19]. Studies have shown that radical abscess incision has a certain effect of reducing the postoperative anal fistula rate and recurrence with no significant damage to the postoperative anal function of patients [20]. However, a meta-analysis involving six studies and a total of 479 subjects showed that, compared with simple incision and drainage, directly opening the suspected fistula significantly reduced recurrence and anal fistula formation (relative risk [RR]=0.13, confidence interval [CI]=0.07–0.24, P=0.38), although the risk of fecal incontinence was significantly increased (RR=3.06, CI=0.7–13.34, P=0.14) [21]. In addition, 34–50% of perianal abscess patients do not form an anal fistula after simple incision and drainage, which implies that these patients can be completely cured through simple incision and drainage without the risk of sphincter injury [22]. Therefore, radical abscess incision of perianal abscess is a controversial issue in the clinical setting. The goal of our treatment is to remove the origin of infection and perform complete drainage without damaging the anal sphincter, in order to reduce fistula formation and abscess recurrence.

The anal gland system is composed of glands, ducts, and openings, and has a flask-like shape. The anal gland is opened in the anal recess and is located between the sphincters [23]. Most prior studies suggested that anal gland infection is the initiating factor in perianal abscess [24–26]. The abscess may extend in three ways (Figure 3 [19]): (1) downward spreading, forming perianal subcutaneous abscess or intersphincter abscess; (2) spreading to the posterior side, forming external sphincter abscess (ischiorectal abscess, presacral abscess); and (3) upward spreading, forming pelvirectal abscess or high intermuscular abscess [27]. Therefore, perianal abscess may occur in the above three cavities at the same time. If these cavities cannot be fully opened and drained, the remnant abscess may cause repeated infection and lead to the formation of anal fistula. TCC is based on the theory of anal gland infection and aims to completely retain the sphincter and to fully drain the infected cavity, in order to reduce postoperative anal fistula formation and abscess recurrence [14].

**FIGURE 3.**
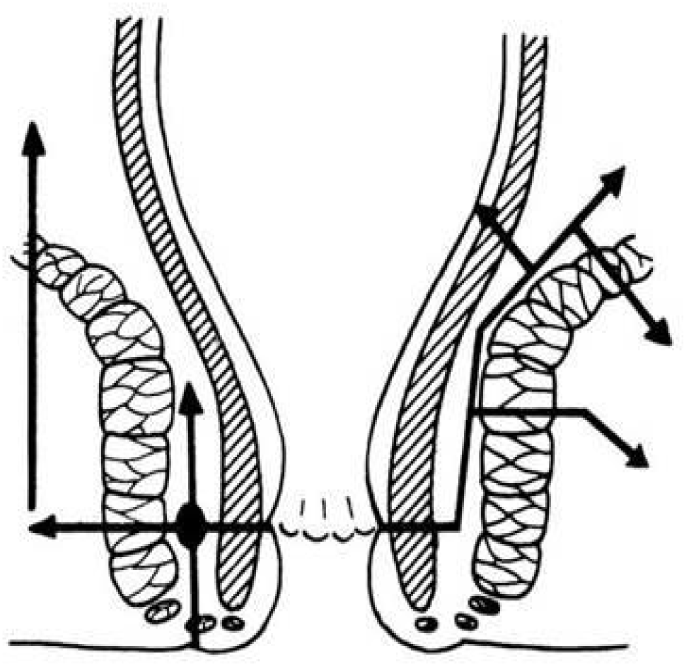
Directions of perianal abscess spreading

The present study found no statistically significant difference in general information between the two groups, which ruled out the influence of individual differences on the study results. The anal fistula formation rate and abscess recurrence rate were 6.2% and 18.0%, respectively, in the TCC group and 1.9% and 8.1%, respectively, in the control group, with statistically significant differences, which suggested that TCC can reduce postoperative anal fistula formation and abscess recurrence. In addition, the anal fistula rate in the TCC group in this multicentre randomized controlled study was similar to that in a previous prospective study (6%), but lower than the rate in a previous retrospective study (13%). The anal fistula rate in the control group was also lower than that in the prospective (34%) and retrospective (39.1%) studies [13, 14]. We presumed that the reason for these discrepancies may be that, in the retrospective study, patients with postoperative anal fistula mainly had pelvirectal abscess complicated with Crohn’s disease or caused by long-term use of steroids. However, TCC is based on the theory of anal gland infection, which suggests that it is a more applicable treatment for cryptoglandular perianal abscess. Moreover, the success rate of simple incision and drainage for non-cryptoglandular perianal abscess is not high. In terms of postoperative pain, the VAS score in the TCC group on day 3 was obviously lower than that in the control group. There was no significant difference in the postoperative VAS score between the two groups on days 0, 1, 2, and 7; however, compared with the control group, the TCC group showed a downward trend in VAS score, indicating that TCC does not increase postoperative pain and is more likely to be accepted by patients. However, rectal non-steroid anti-inflammatory drugs(NSAID) are routinely used in our hospital to relieve postoperative pain. In a previous retrospective study, the postoperative VAS score on day 7 in the radical abscess incision group with stringing was 6.5±1.3 points, which was significantly higher than that in the other two groups [12], and also significantly higher than that in the TCC group (1.9±1.2 points) and the control group (2.2±1.1 points) in the present study. It is further suggested that TCC allows complete sphincter retention and provides postoperative pain relief. The hospitalization duration was significantly lower in the TCC group than in the control group, and there was no significant difference in wound healing time between the two groups. However, the healing time of the TCC group showed a tendency to decrease, which means that TCC can cure the disease without increasing the risk of anal sphincter damage and the healing time, and has no impact on the quality of life of patients. The comparison of postoperative anal function between the two groups showed no cases of anal incontinence, indicating that TCC would not further lead to loss of anal function. The success rate in the TCC group was significantly higher than that in the control group, which further confirmed this inference.

In conclusion, this randomized controlled study showed that TCC for perianal abscess charaterized by reduced postoperative anal fistula rate, reduced abscess recurrence, increased success rate, no anal sphincter injury, and reduced postoperative pain. It is a safe and effective method for the management of cryptoglandular perianal abscess.

## Data Availability

the availability of all data referred to in the manuscript

